# Virus evolution affected early COVID-19 spread

**DOI:** 10.1101/2020.09.29.20202416

**Authors:** Derek Corcoran, Mark C. Urban, Jill Wegrzyn, Cory Merow

## Abstract

As the SARS-Cov-2 virus spreads around the world afflicting millions of people, it has undergone divergent genetic mutations. Although most of these mutations are expected to be inconsequential, some mutations in the spike protein structure have been hypothesized to affect the critical stage at which the virus invades human cells, which could affect transmission probability and disease expression. If true, then we expect an increased growth rate of reported COVID-19 cases in regions dominated by viruses with these altered proteins. We modeled early global infection dynamics based on clade assignment along with other demographic and meteorological factors previously found to be important. Clade, but not variant D614G which has been associated with increased viral load, enhanced our ability to describe early COVID-19 growth dynamics. Including clade identity in models significantly improved predictions over earlier work based only on weather and demographic variables. In particular, higher proportions of clade 19A and 19B were negatively correlated with COVID-19 growth rate, whereas higher proportions of 20A and 20C were positively correlated with growth rate. A strong interaction between the prevalence of clade 20C and relative humidity suggests that the impact of clade identity might be more important when coupled with certain weather conditions. In particular, 20C an 20A generate the highest growth rates when coupled with low humidity. Projections based on data through April 2020 suggest that, without intervention, COVID-19 has the potential to grow more quickly in regions dominated by the 20A and 20C clades, including most of South and North America.

## Introduction

Novel Coronavirus Disease 2019 (COVID-19) is causing widespread morbidity and mortality globally [1,2]. The SARS-Cov-2 virus responsible for this disease has now infected 22.5 million people through August 2020 [3]. As the virus has spread around the world, it has mutated into divergent clades with different prevalence in different geographic regions [4,5], although all five of the clades described today are circulating around the globe. Clades known as 19A and 19B emerged in Wuhan and clade 20A emerged from 19A and was prominent in the European outbreak in March. Both 20B and 20C are considered subclades of 20A [6]. To date, we do not know if these geographically dispersed clades are associated with altered disease dynamics. Yet, the models used to inform interventions, travel restrictions, and health care capacity generally assume equivalent pathogenicity and transmission potential [7]. Understanding if genetic clades differ in their infectivity and, if so, where they are most prevalent would aid efforts to design effective intervention strategies that control the virus and end the pandemic.

One of the greatest uncertainties for projecting future COVID-19 risk is how its evolution will affect its future transmission dynamics. Although most mutations are expected to be neutral and thus would not alter viral transmission or infection dynamics, the chance exists that one or more mutations affect viral traits that can either increase or decrease the manifestation of the disease in humans. For instance, the flu strain that caused the deadly 1918 pandemic eventually evolved into a less virulent form that still circulates today [8]. In contrast, the bubonic plague evolved into a form with airborne transmission that enhanced outbreaks [9]. The evolution of virulence often trades off against transmission rate, thus offering two pathways for adaptive evolution, with the exact course taken dependent on multiple, interacting factors [10,11].

SARS-Cov-2 might be particularly likely to evolve given its high prevalence and global distribution [12]. Early reports indicate possible differences in growth rate and viral loads associated with certain mutations [13]. Although broad genomic divergence might be associated with functional changes in the virus, we pay particular attention to the evolution of the outer spike proteins of SARS-Cov-2. The evolution of the outer spike glycoproteins of SARS-Cov-2 may influence infection rates because these structural proteins bind the virus to host cells via the angiotensin-converting enzyme 2 (ACE2), which ultimately allows the virus to bind to and enter the host’s respiratory cells and thereby cause COVID-19 [14]. This prediction is well-supported by theory, electron microscopy, and experiments [7,15,16] as well as observed mutations in spike proteins causing enhanced virulence in other coronaviruses [17]. The evolution of SARS-Cov-2, likely in an intermediate host between bats and humans, led to its spread in humans [18]. Further evolution of these spike proteins could enhance binding to human ACE2, enhancing transmission, infection rate, virulence, while exacerbating symptoms. Specific interest has focused on codon 614 of an especially glycosylated region of the viral spike protein, where positive natural selection has been confirmed for the D614G variant [7,12,19].

Here, we estimate whether clade and the D614G variant, when combined with weather and demographics, affected COVID-19 growth rates early in the pandemic spread. We examine early growth before widespread intervention was prevalent (up to April 13, 2020) because intervention altered the ability to separate biological mechanisms such as evolution from regulated human behaviors. We build on a previous model that successfully described global infection dynamics early in the pandemic and highlighted effects from ultraviolet light (hereafter UV light), temperature, humidity, and age structure on COVID-19 growth rates [20]. In this study, we modeled the growth rate of COVID-19 as a function of these same variables and one of two possible ways to describe virus evolution: variants of codon 614 or genetically distinct major clades. The 614G variant has increased in frequency around the world [13], and studies suggest that it might be more infectious than its predecessor, 614D, in experiments performed within cell lines [13,21]. However, it is unclear if infected individuals carrying this mutation are more contagious and there is no published support that 614G is more virulent [22] Hence we tested whether a higher prevalence of 614G variants within a population was associated with a higher growth rate of the disease. Additionally, since the clade distinctions summarize major genetically distinct groups (encompassing numerous variants) that achieve significant frequency and spread (∼20%), we also tested whether any of these clades were associated with increased growth rates. It should be noted that D614G does not define the clade divisions though it is considered the predominant missense mutation distinguishing isolates originating in Asia from those in Europe [13]. Several other variants are used to distinguish the clades in the 29.9 Kb genome. Clades 19A and 19B are associated with C8782T and T28144C, respectively. 20A has variants C3037T, C14408T, A23403G. 20B is associated with G28881A, G28882A and G28883C, and 20C contains C1059T and G25563T [23]. We compared six different models using boosted regression trees [24,25]: (1) a null model (intercept only), (2) weather and demography, (3) clades, (4) variants, (5) weather, demography and clades, and (6) weather, demography and variants. The weather and demography model was demonstrated to successfully predict COVID-19 growth rates in [20] using a different approach (linear regression). Boosted regression trees have several advantages over traditional regression approaches, including that this approach includes interactions when appropriate without the need to specify them, removing variables that are irrelevant, and being able to adjust to any response shape [24–26].

We modeled the maximum growth rate of COVID-19 cases to restrict analyses to the early growth phase before social interventions reduced transmission, but after community transmission began, and when most people were still susceptible to this novel virus. We used growth rates in contrast to total cases because they are less sensitive to bias as shown in [20]. The analysis was restricted to political units with >40 cases to eliminate periods before local community transmission. These decisions resulted in data from 79 countries and 33 states or provinces where genetic data were also available. We restricted the sample to the three worst one-week intervals in each polity separately to characterize maximum potential growth rates.

## Results

The best performing model included weather, demography and clade based on both training and independent testing data, such that the root mean square error (RMSE) was 25% lower and explained 15% more variance than the next best model. The model including weather, demography and variant did not substantially improve descriptions of disease growth relative to using only weather and demography (Figure 1). For more detailed results check on Figures S3 and s4 in supplementary materials.

**Figure 1.**
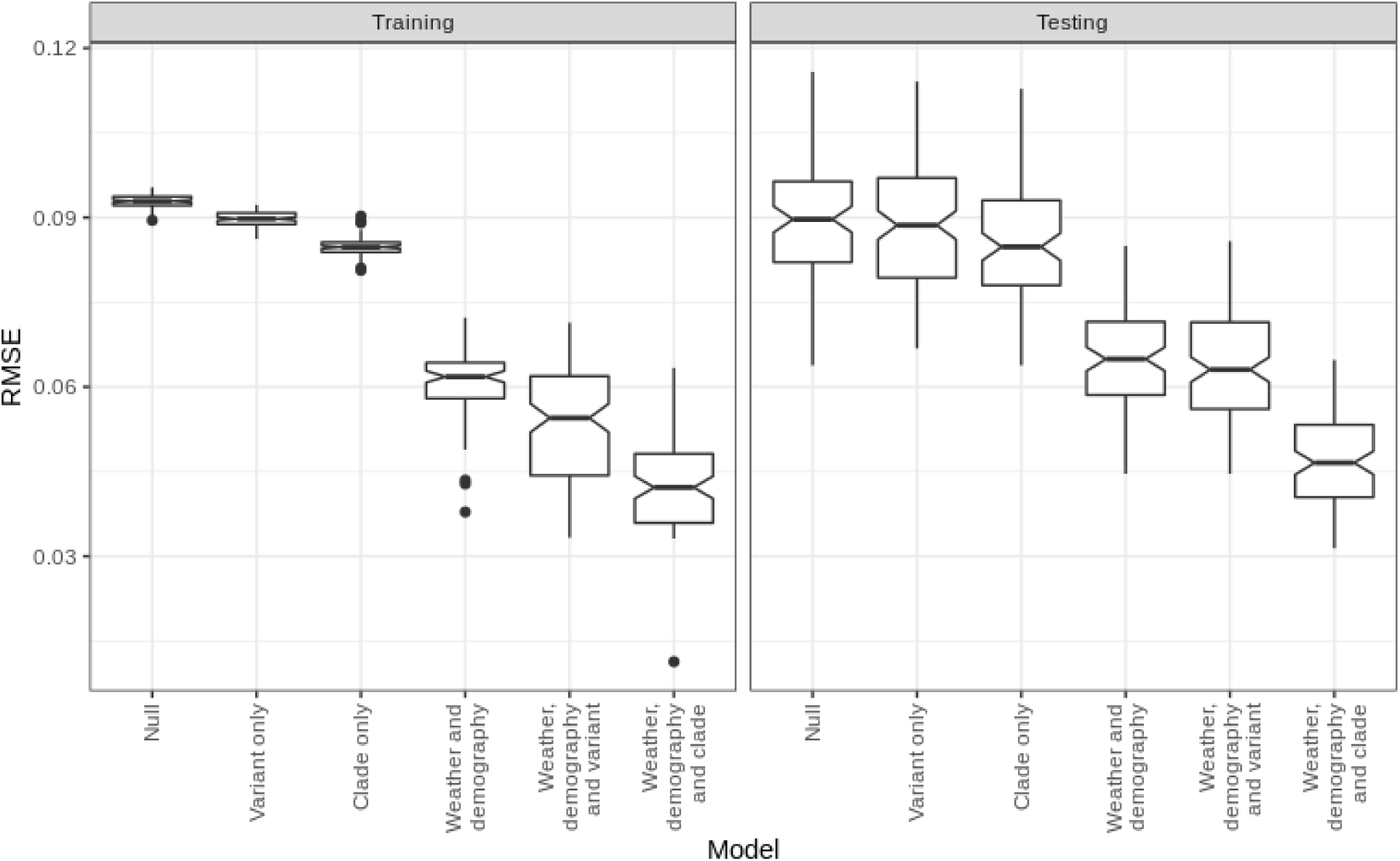
Boxplot of the root mean square error (RMSE) of the models fitted for the train and test sets, where the notches in the boxplot represent the 95% confidence interval of the median and dots indicate outliers. Smaller RMSE indicate better models. In both the training and test sets, the best performing model is the weather, demography and clade model. The variant, demography and weather model on the other hand does not improve on the weather-only model in the test set, which suggests that the variant is less informative than clade.

We next assessed the importance of each factor in the model, which describes the reduction of squared error attributable to each variable. For all models that include the weather data, UV light was the most important variable, where COVID-19 growth rate diminishes as UV light increases. However, the summed importance of all clades was higher than for UV light, suggesting that the collective importance of clade prevalence is comparable to that of the dominant weather factor (UV) (Figure 2).

**Figure 2.**
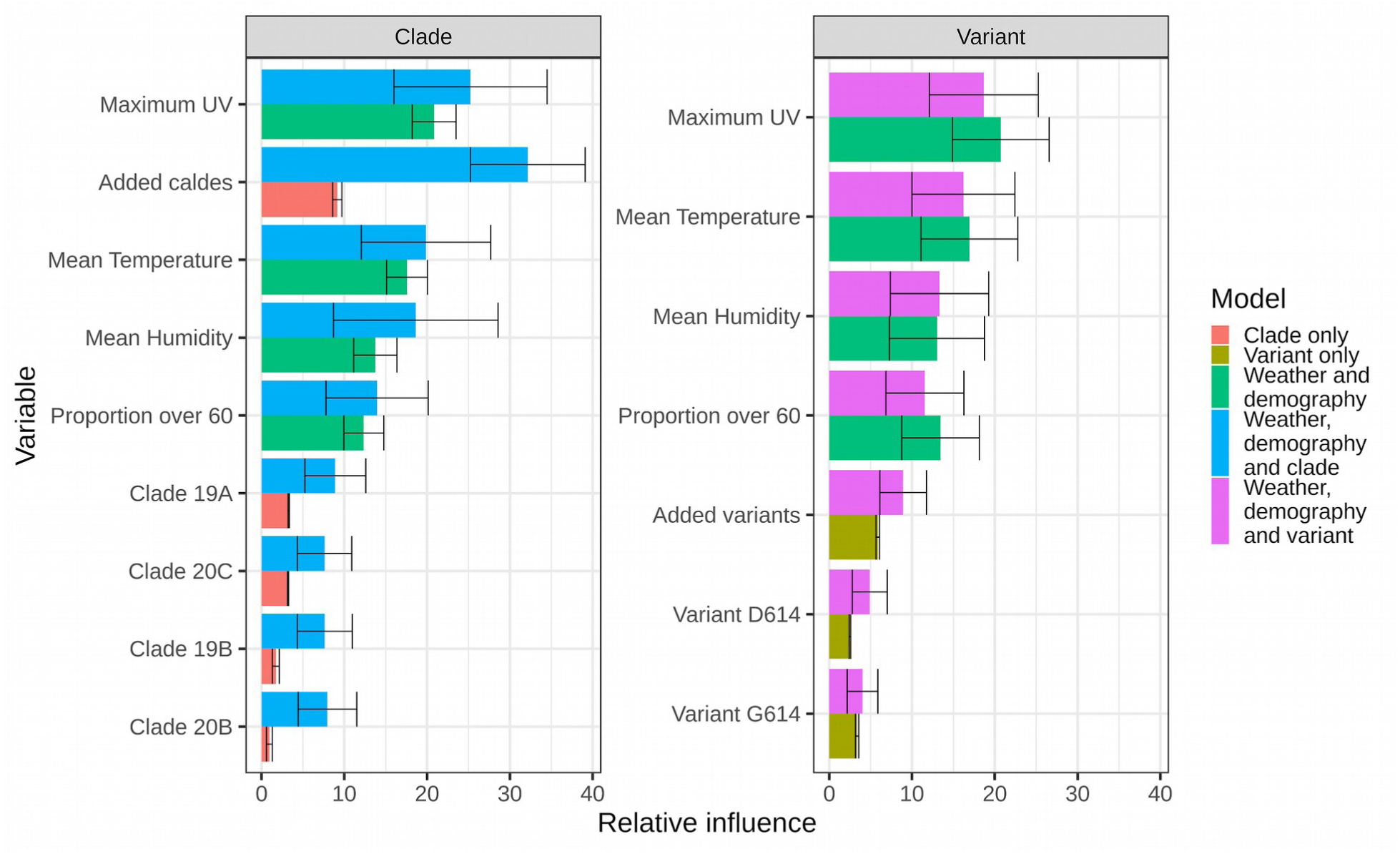
Relative influence of variables for all models. Error bars indicate 95% confidence interval. UV light is the most important variable in all models that contain that variable, however the summed importance of all clades in the weather, demography and mutation model is more important than UV light.

Clade prevalence had higher importance when included in models that also included weather which indicates interactions among these variables. We calculated Friedman’s H-statistic [24,25] to assess the strength of interactions [25] between clades and other weather and demographic variables, which ranges between 0 and 1, with larger values indicating stronger interactions (supplementary table 1). Among the clades associated with higher growth rates, 20C has the highest interaction with relative humidity (H=0.27, Figure s3), almost double the interaction strength of the next highest interaction. Low relative humidity, combined with a high proportion of 20C, was associated with higher COVID-19 growth rates. In experimental and correlative studies, low humidity contributes to the spread of several respiratory viruses [20,27,28]. This interaction suggests that low humidity might enhance the spread of the more infectious clades of SARS-Cov-2.

Response curves indicate how COVID-19 growth rates depend marginally on clade prevalence (Figure 3). Higher prevalence of clades 20A and 20C were associated with higher growth rate, whereas higher prevalence of clades 19A and 19B were associated with decreased growth rates. COVID-19 growth rate did not consistently vary with the prevalence of clade 20B. These patterns were consistent with those found when applying classic linear regression analyses (Figure 3), ensuring that trends we detected were not an artefact of the machine learning algorithm.

**Figure 3.**
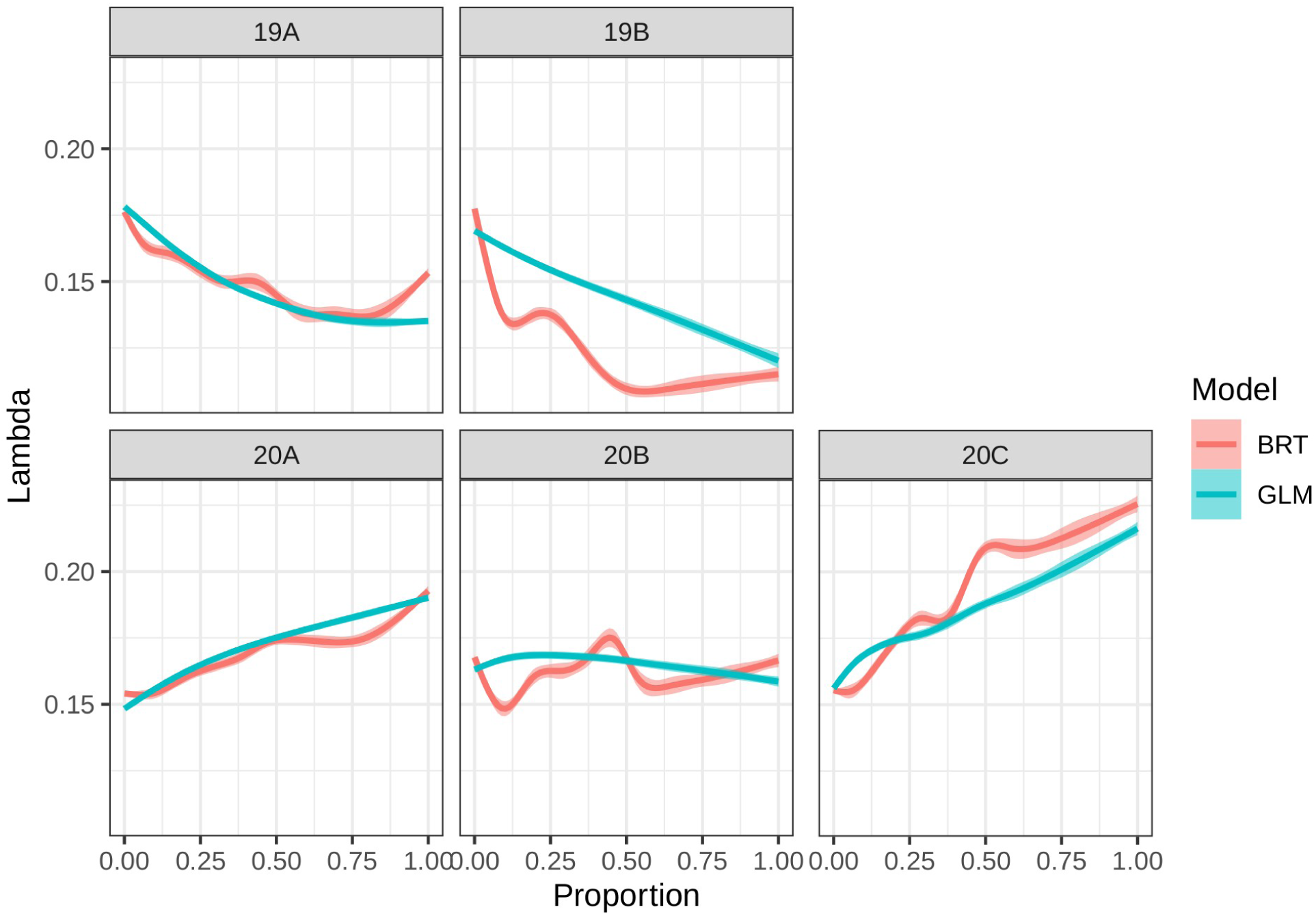
The Boosted Regression Trees (BRT) model shows that COVID-19 growth rate increases with the proportion of clades 20A and 20C, does not have a strong association with the proportion of clade 20B, and decreases with the proportion of clades 19A and 19B. The same patterns hold when applying an alternative generalized linear model (GLM). These response curves are the result of the smoothed prediction of both types of models (using GAM) over 8,000 simulated points that maintain accurate summed proportions (see methods).

In order to validate the models built on data collected before mid-April, we tested predictions against independent data for late April and May 2020. We calculated clade prevalence, average weekly growth rates, and 14-day lagged weather conditions, assuming the proportion of people over 60 remained constant over this short period following methods in [20] and compared the two best performing models (weather, demography and clade vs. weather and demography) based on RMSE. Although widespread health interventions were already becoming common during this time, thereby reducing growth rates compared to the time period when our model was fit, the weather, demography and clade model still made better predictions than the weather-only model in both April and May (RMSE = 0.106 versus 0.112 in April and RMSE= 0.136 versus 0.148 in May). RMSE during April and May was larger than RMSE during cross-validation (based on February -early April data), consistent with findings in [20], likely due to increased intervention during this time period, leading to consistently overestimated growth rates.

From January to September, clades 19A and 19B decreased from 96.7% of the 156 total cases tallied in January to 5.2% of the 232 cases tallied in August, and 11.7% of the 51 cases analyzed so far in September (Figure 4). The three clades in group 20 all increased in proportion relative to clade 19. The most rapid increase in prevalence was for clades 20A and 20B. Clade 20C increased rapidly initially in prevalence but has since leveled out at the global scale but may still be increasing in North America. Given our results, we expected clades 20A and 20C to increase, if they are representative of total virus distributions worldwide. In August, over 60% of the cases registered in the world were classified as clade 20A. However, the change in proportion of these clades has not followed this pattern in various regions of the world. Using a binomial generalized linear model of changes in clade proportions through time, we found significant interactions of clade, date, and region (Null deviance: 12,415.4 on 974 degrees of freedom; Residual deviance: 2,152.3 on 915 degrees of freedom, significantly different from null model with Chi square p-value less than 0.0001). For both North America and South America, clade 20C grew to the highest proportion, with 55% in North America and 33% in South America. In line with growth rate model results, these two continents have had the highest total number of cases. At the same time, the peak monthly prevalence for the 20C clade in Europe was in March, which coincides with the highest growth rates in Spain and Italy, two of the countries with the largest number of cases in Europe.

**Figure 4.**
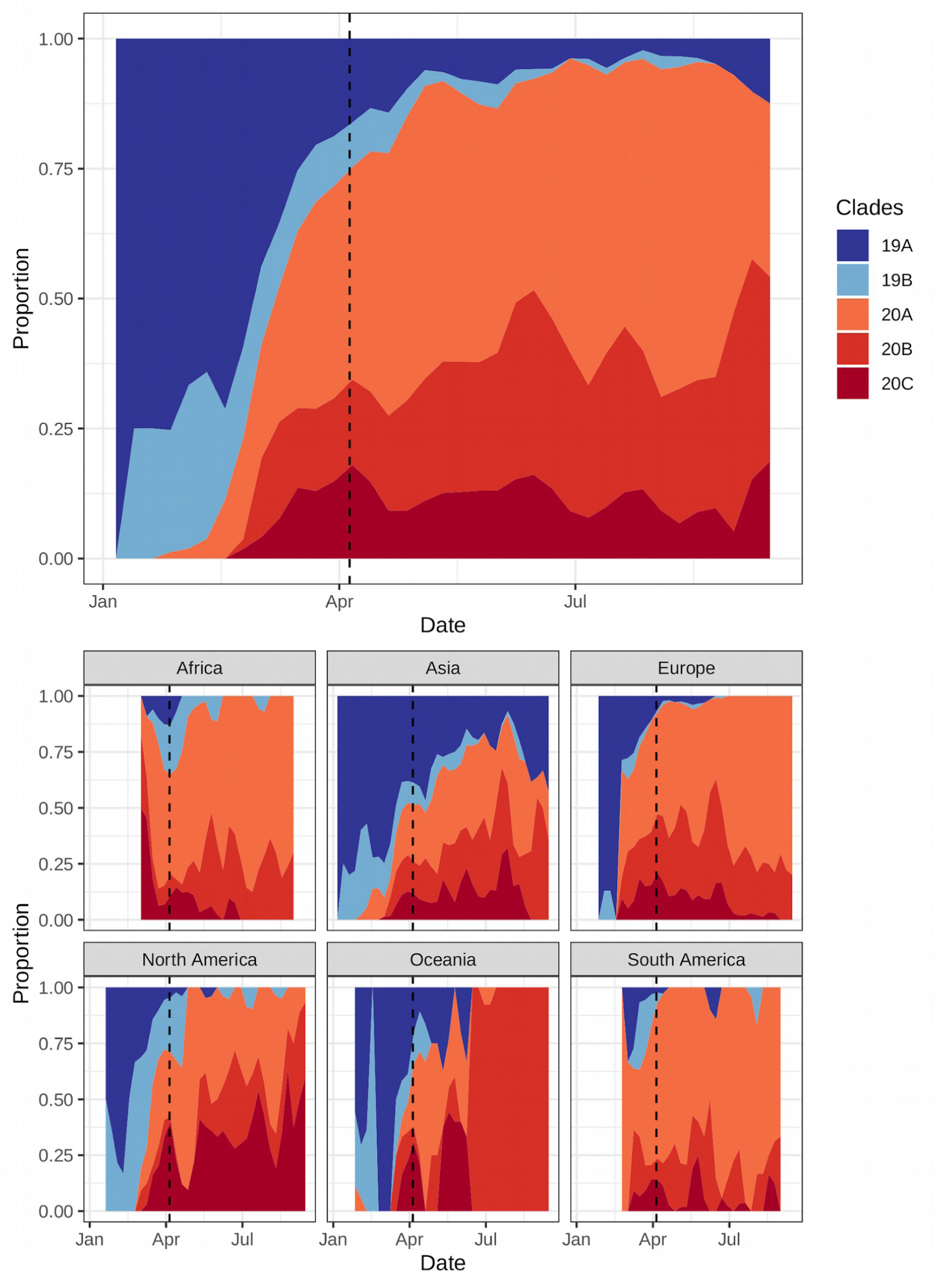
Proportions of each clade globally (top) and by region (bottom). Clade 19 (blues) diminished over time, while clade 20 (reds) increased during that same period. Among the type 20 clades, 20A and 20B have increased to similar proportions globally while 20C had more moderate proportional growth. However, clade 20C has increased most dramatically in South and North America, two regions where COVID-19 has subsequently grown quickly. The dashed line represents the last date used for fitting the models.

During the month of August, which is our last full month for which data was available, either clade 20A or 20C was the most abundant clade in several regions:Iin South America, 81% of the cases in August were from those two clades, followed by 78% of the cases in Europe and Africa, and 75% of the cases in North America. In contrast, all of the cases in Oceania were classified as 20B, a clade associated with lower growth rates according to our results.

## Discussion

We provide new evidence for an evolutionary effect on the COVID-19 growth rate early in the pandemic. Although two different measures of SARS-Cov-2 evolutionary divergence were tested, only the prevalence of different genetic clades improved the explanatory power of COVID-19 growth rate before widespread interventions were implemented and predictions after intervention. Although UV light was still the single most important predictive factor, cumulative importance across all five major clades was comparable to the importance of UV light, indicating that mutations resulting in the formation of major clades might have been a critical factor shaping early spread. Both clades 20A and 20C were associated with higher growth rates while clades 19A and 19B were associated with lower growth rates. Notably, a considerable amount of variation in growth rate remained unexplained, which we attribute to variation in social behavior and health care availability as well as more comprehensive intervention, testing, and care policies that were implemented at varying times depending on polity. The three continental areas with the highest proportions of either clade 20A and 20C during the month of August 2020 were South America, Africa and North America, suggesting the potential for higher growth in these regions in future time periods.

It is notable that mutation D614G did not improve model performance per se, whereas finer distinctions among clades -the variants discussed about -that included this mutation did improve model performance. Variant 614G maps to all genomes in clade 20 (Supplementary Table 2), and even when there is not a one to one concordance between variants and clades, 99.65% of the cases that belong to the 614G variant, are part of the 20X clade with only 0.35% remaining being part of the 19X clades, of these, 45.43% of the cases belong to the 20A clade, 36.76% to 20B and 17.45% to 20C.

However, among these clades, only 20A and 20C were associated with increased growth rate while clade 20B was not (Figure 3). Consequently, clade 20B’s patterns confound any directional trend when combined with patterns for clades 20A and 20C, making variant D614G appear to have low explanatory power. Hence, we found that variant D614G is important, however only particular clades that include this mutation appear to be associated with increased COVID-19 growth. While clinical studies have associated the spike protein D614G variant with increased viral load, the full impact of D614G is not conclusive in terms of increased transmission or severity [22,29]. It is difficult to determine whether specific variants are neutral and increasing from demographic processes, or in fact, increasing the rate of transmission and/or severity. Van Dorp and colleagues [30] examined approximately 200 homoplasic variants across over 45,000 genomes and did not find any relationship between transmissibility and 614G. However, a study focused on county level models in the United States examined the impact of D614G in conjunction with population density and found that the presence of 614G significantly increased transmission [31]. The data suggests that the path to developing mitigation policies and therapeutic tools will need to encompass a broader view of the role of these clades. The prevalence of clades 20A and 20C is acknowledged in recent studies and, with it, an associated focus on finding rare or descendent variants outside of D614G that may contribute to this [32,33].

Another interesting aspect of our research is the finding that environment and viral characteristics can interact in complex ways that might enhance or diminish the infectivity of a viral clade. At high humidities (approximately 80%), we might not see any difference in the growth rate of the disease. In contrast, at low humidities, the genetic composition of SARS-Cov-2 could become more important. Few studies combine both an understanding of the evolution of disease agents in combination with weather or climate variables. However, many other such interactions are likely when we begin to explore these two factors in disease dynamics more fully.

Some limitations of our study are important to recognize when interpreting the results. First, genomic information remains limited, and although we analyzed 9000+ genomes, sample size issues remain important (6). In particular, the number of cases available for the estimation of the proportion of each clade or variant in a given polity at a particular point in time was sometimes low. The number of cases within a polity ranged from 1 to 110, with a median of 21. However, we performed a sensitivity analysis, removing polities with fewer than five cases, and demonstrated that removing polities with low numbers of samples resulted in qualitatively similar results, with the exception of a flatter response for clade 19B (Figure S5). To further test our analysis, we developed another, sensitivity analysis, we generated 95% multinomial confidence intervals for the clades proportions following <u>(Sison and Glaz 1995)</u> and made 100 replications of analysis selecting randomly for each polity the lower upper or estimate, and generated a new analysis with that. As was the case with the prior sensitivity analysis, the results were very similar with clade 19B again being the only one differing by having a flatter response (Figure S7). A related caveat is that the flexibility of BRTs allows for the possibility of overfitting to potentially idiosyncratic trends in clade prevalence. To address overfitting, we took two steps. First, we used replicated, balanced cross-validation to produce an ensemble of models, and weighted these models by their predictive performance on withheld data to make our final predictive model. Second, we also fit simpler generalized linear models that showed the same qualitative patterns (Figure. 3). Although we find a correlation between COVID-19 growth rate and the prevalence of certain clades, we cannot yet make conclusive statements about the causality. The chance remains, as with all correlative models, that these correlations are spurious and reflect spatial structure in neutral evolution that happen to be also associated with outbreaks. For instance, the high growth rates of COVID-19 in North and South America could have been aided in part by a more rapidly spreading viral variant, or alternatively the growth could have occurred for a number of reasons and this variant just happened to have been prevalent there. Only through controlled trials could we begin to separate out causality from correlation, and given the inability to do this, we can only rely on such observational evidence in combination with future laboratory studies to suggest patterns that merit further detailed study.

Despite these caveats, results suggest multiple key pathways for further monitoring and testing. First, we found that evolution contributed to variation in initial growth rate among political units as much as UV light. Our results from real-world observations of transmission and disease growth suggest that the current genetic clades matter more than the widely studied D614G mutation. We did not find evidence that the mutation D614G alone contributed to early growth rate, in spite of findings elsewhere that this mutation increased more readily in clinical settings and appeared to be under positive selection [12,19,22]. Continued genomic monitoring is critical to detect whether certain clades associated with the current, or that arise de novo, change in frequency over time.

Although SARS-Cov-2 is unlikely to evolve as quickly as influenza, our results demonstrate an important potential for existing genetic changes to already have influenced disease growth rate. Additional research is needed to test if different clades vary in the types or severity of COVID-19 symptoms -the increase in growth rate is not sufficient to imply that clades 20A and 20C are more virulent. The opposite could be true if the virus evolves enhanced transmission at the cost of its virulence. On the other hand, if reporting is biased toward the most symptomatic cases, then these clades might be expressed more severely rather than spread more rapidly. Correlating infected clade with symptom severity in an unbiased sample is a critical research need.

Importantly, a large amount of unexplained variation suggests that social behavior and public health interventions likely contribute much more to variation in COVID-19 spread than evolution or weather [34]. Moreover, the large pool of uninfected hosts will continue to dominate the epidemiology of this disease. However, evolutionary changes and weather could provide a greater or lesser potential for rapid spread especially once social interventions are relaxed or once a smaller pool of people are susceptible either through exposure or vaccination. The dominance of the genetic clades associated with high growth rate in regions that experienced rapid growth of COVID-19 in later months is suggestive, but not yet conclusive, that these clades have the potential to influence COVID-19 dynamics.

Modern epidemiology can anticipate many features of disease outbreaks and mitigate their effects on human health through myriad public social, health, and pharmaceutical interventions. Yet, we often still cannot predict with certainty how a particular agent will evolve and whether these adaptations will enhance or reduce virulence or transmission. As we understand more about the evolution of disease agents, such as SARS-Cov-2, this information should aid our ability to make accurate predictions about future outbreaks and design effective public health interventions in order to end this pandemic and prevent new ones from emerging.

## Methods

### COVID-19 dataset

Maximum growth rates of COVID-19 cases were modeled to limit research to the early growth period before transmission was decreased by social measures, but after transmission to the population started and when most people were still susceptible to this new virus. The average maximum growth rate (*λ*) was calculated as the exponential increase in cases: ln(Nt) – ln(N0)/t, where N*t* = cases at time, *t*, and N0 = initial cases. We used a repeated measures design for the three worst one-week intervals in each political unit (country or state/province depending on available data [3]), where *t* = 7 days (see sensitivity analysis in [20] which demonstrated that other time windows from 1-7 days lead to similar results due to the temporal autocorrelation of weather). There is considerable variation in testing and reporting of COVID-19 between countries and even smaller political units which makes models based on count data unreliable. Comparatively, using growth rates should remain resilient to biases introduced by differences in report rate, assuming that detection probabilities do not change substantially in a given polity during the short, one-week period. We limited analyses to polities with >40 cases to ensure that the transmission of the disease was local. which led to a database comprising 128 countries and 98 states or provinces.

We obtained daily infection data from [3] and 3-hour weather data from the ERA5 reanalysis for the 14 days preceding case counts [35]. We averaged these values to reflect the possibility that infection could have occurred during the previous 14 days, consistent with the 1-14 day infective period widely reported [36]. Given the uncertainty in the joint distributions of symptom onset, testing, and reporting, as well as not knowing the degree to which variables influenced COVID-19 case growth via transmission versus the expression of symptoms (e.g., vitamin D immune function), we chose to average across the potential period of infectivity, thereby assuming weather each day in the preceding 14 days was equally important. However, results were robust to a range of other assumptions when calculating lagged weather variables, including weighted means centered on 6, 9, and 12 days as well as different variances in [20]. We used fine-scaled weather data rather than long-term climatic monthly means to model observed weather-outbreak dynamics. Weather data was weighted by population size in each 0.25° grid cell within each political unit to capture the weather most closely associated with outbreaks in population centers.

## Genomic clade dataset

We evaluate how well the proportion of each of the five major clades as defined by NextStrain, and the proportion of known mutations of codon 614, maximum COVID-19 growth rate across political units (6). These proportions were based on the observed proportions of clades and the variant scored in each political unit (sensitivity analysis based on upper/lower bounds of these multinomial proportions yielded similar results; Figure S7, where only 19B seems to have a flatter response than our results). We used 319 estimations of the proportions for 108 political units comprising 9,001 cases of COVID-19 for clade proportions, and 317 estimations of the mutation proportions for 107 political units (only Ghana is missing) comprising 7,457 cases. The number of cases for the estimation of proportions of clades varied among territories, where we estimated the proportion using a minimum of 1 individual case for 20 territories and a maximum of 110 individuals for three territories, a sensitivity analysis was made using only the polities that had at least 5 cases to calculate the proportions. To see how small samples sizes affected results, we performed an analysis in which we eliminated data from polities with fewer than five genomes. Results were qualitatively similar to those using the full dataset, except that clade 19 had a flatter response (see Figure S5). In the case of the codon variants, we had data for 29 polities with a single case and three with a maximum of 94 cases. For each polity, we selected the worst three one-week intervals of disease growth rate when available following [20], however, there were 5 polities where only 2 weeks were available for the time-frame selected for clades, among those was Ghana, which was not part of the dataset for mutations. Within the polities that had all 3 weeks, there were differences of a minimum of 14 days between the first and last date, and a maximum of 56 days. The earliest date used was January 22 for one territory, and the latest estimation was from April 05 for 1 territory. The most common clade for each polity is shown in Figure 5, and the proportion of each clade for each territory is shown in Figure s6.

**Figure 5.**
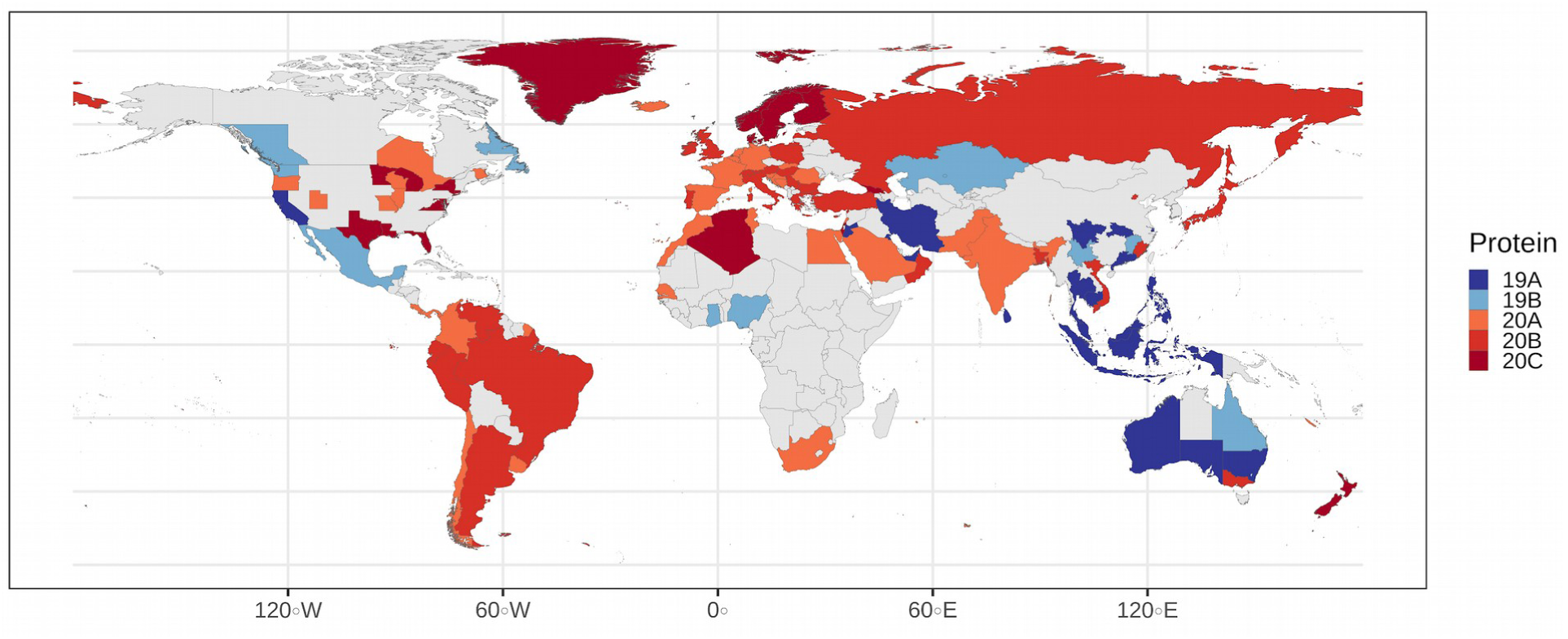
Most common clade on each polity as seen in the map, clades 19A and 19B dominate in Oceania and southeast Asia, whereas clades 20A, 20B and 20C dominate in South America, Europe and North America.

We used cross-validation to estimate relationships between explanatory factors and COVID-19 growth rate, which uses the performance of models trained on a subset of data and applied to left out data. Such methods are especially important to avoid overfitting more flexible models. We applied stratified 10-fold cross validation, repeated five times, and trained models on 90% of the data and tested them with the remaining 10% based on a constrained randomization procedure to generate the folds. We stratified by polity such that if one week was assigned to the training set, the rest of the weeks were also assigned to the training set to ensure that information about a polity in the test data was not also included in training data, thus potentially augmenting confidence in the model. Given this procedure, not every split was exactly 90:10, so we also constrained the randomization to be < 5 cases from an exact 90:10 split, which produced 95 different training sets and 95 different testing sets for the clades. We repeated the same process for the variant, and this procedure yielded 121 different training and testing sets.

We used boosted regression trees (BRTs) to fit models because they offer several advantages over other regression techniques. BRTs are a type of machine learning technique that seeks to optimize the predictive accuracy of out-of-sample data in an iterative process and using an ensemble of regression trees. By focusing on prediction, BRTs can provide a better estimate of predictive accuracy in contrast to traditional generalized linear models (e.g., linear regression). They tend to avoid including irrelevant variables, and interactions between variables are inherently included without the need to specify them a priori [24,37]. Furthermore BRTs can fit any shape of response, and hence avoid the possibility of underfitting. However, due to this flexibility, cross validation is necessary in order to avoid overfitting. Since BRTs depend on tree-based methods, the number of bifurcations of each variable together with the reduction of the residual error in each of those can be used to calculate the relative influence of each variable [24,38].

For each training set we did a 10-fold-5-repeated cross-validation to reduce overfitting, selecting the best model optimizing the value of Root Mean Squared Error (RSME) following [39] using the caret package [40] and boosted regression trees through the *gbm* package [38]. This process was done 95 times for the clades and 121 times for the mutant, i.e., once for each training set. For the hyper-parameter tuning, we used the following grid for the interaction depth 1, 5, 9; the number of trees for boosting were 1, 50, and a sequence 50 by 50 up to 1500 trees; shrinkage of 0.1 and 0.01 and a minimum number of observations in the terminal nodes of either 10 or 20.

We next created a weighted ensemble model that best predicted the withheld data in each of the 95 different training and testing sets. This was done in order to further diminish over-fitting by then weighting the value of each fitted model by the inverse of our overfitting metric. The overfitting metric was based on the cubic root of the difference of the R-squared value in the test and training sets. We chose the cubic root to give high weight to models where the training and testing performance was the most similar, and thus to reduce overfitting even more than in most applications because our aim was to predict using the simplest models supported by the data. At the same time, we calculated the weighted testing model performance as the root mean squared error (RMSE) and variance explained (R^2) for each model (except the null model), thus resulting on a 95 and 121 testing metrics, to understand how sensitive the ensemble model was to different test sets.

Given the flexibility of BRTs, we also applied traditional generalized linear regressions to the same data to see if results were robust to methods. To be consistent with the BRT models, we used an ensemble of the best performing GLMs. For the GLM, we fitted the weather and protein model with all the variables, including linear and quadratic terms for every variable. We next assessed model performance using the Akaike information criterion, and then assessed further models of a similar structure but without two of the highly correlated variables. From our model set, we averaged the coefficients for the ensemble of models that summed to 0.95 in Akaike weights. Thus, the GLM models also developed a set of predictions based on a model-averaged ensemble of predictions, and thus accomodate more flexible curvilinear relationships with explanatory factors as compared to the single best polynomial model.

Results from GLM and BRT approaches were highly similar (Figure 3), suggesting concordance, a lack of overfitting for the BRTs, and qualitatively similar results, regardless of which approach is applied. We focus on the BRT results in the main text given the further checks that we applied to prevent aggressive overfitting.

## Models

We generated six different models to test how much of the variance was explained by weather, demography, genetic classification, and/or the presence of the mutation D614G. We first fitted a model that only used weather and demographic variables. Six different models were tested, beginning first with the weather and demography model evaluated previously, where the growth rate for the prior seven days was explained only by weather factors and the proportion of the population with age over 60 years old as it was demonstrated in previous work [20]. In this model, the COVID-19 growth rate (*λ*) for the prior seven days was modeled as a linear function of weather variables calculated over the 14 day interval preceding the estimate of lambda and the proportion of the population over age 60. Weather variables included the mean daily temperature *λ*, the maximum of the mean daily UV, and the mean daily relative humidity. Sensitivity analysis in [20] explored different combinations of these and other variables as well as lagged intervals ranging from three to 21 days and found that predictions were robust to these various possibilities. Hence we use the model selected in that study as the starting point for examining whether additional information on SARS-Cov-2 evolution can improve explanatory power. The second model tested if the same COVID-19 growth rate was explained by the proportion of clades in a polity, without taking into account either weather or demography. Concurrently, a similar model was fitted with the same rationale, but using the D614G variant instead of clade classifications, where we tried to explain the growth rate by the proportion of n −1 of the alternative clade or mutation, leaving one out to ensure model identifiability following the conventions used to analyze multinomial compositional data [41]. The main purpose of this was to test the degree to which these models explained the variation of the growth rate of the disease. Then we built models using the weather and demographic variables with either the protein or mutation information to test if a full model with either classification improved on the weather/demography models. All of these models were then compared to null models (without explanatory factors and just an intercept) to test if any of them were better than just using the mean as an estimated growth rate.

## Response curves

To build the response curves of COVID-19 growth relative to the proportions of each clade (Figure 3, describing the marginal response of growth rate to the prevalence of each clade), we used an approach that ensured that proportions summed to one and maintained correlations between clades to incorporate observed interactions. In order to do that, we used a methodology to generate discrete random variables constrained to marginal probabilities and correlations following Barbiero (28). We used each of the counts of clades in each of the polities to calculate both the correlation and marginal distribution of the frequency of each of the clades, and simulated data sets until the concordance of the correlation between simulated data and real data was less than 0.1 for all clades in 8,000 simulations. For each site, we simulated the frequency of each protein and then calculated the correlation of the proportions in order to compare it to the proportion in the dataset (shown in supplementary Figure 1). We then used these simulations together with the weather and demographic variables fixed to their respective means in order to predict lambda for each simulated data set. This procedure was designed to capture the variability among the simulated datasets. Finally, we smoothed these predicted values of lambda using a generalized additive model (with smoothing determined by maximizing cross-validation). This procedure was used for both the BRT and GLM predictions (Figure 3).

To test for the strength of interactions between variables, we used Friedmen’s H statistic [25], which tests the strength of the interaction between variables on a scale from 0 to 1, where 0 means no interaction, and 1 means total dependence. This statistic was calculated for all pairs of interaction in each model.

## Supplementary material

**Figure S1.**
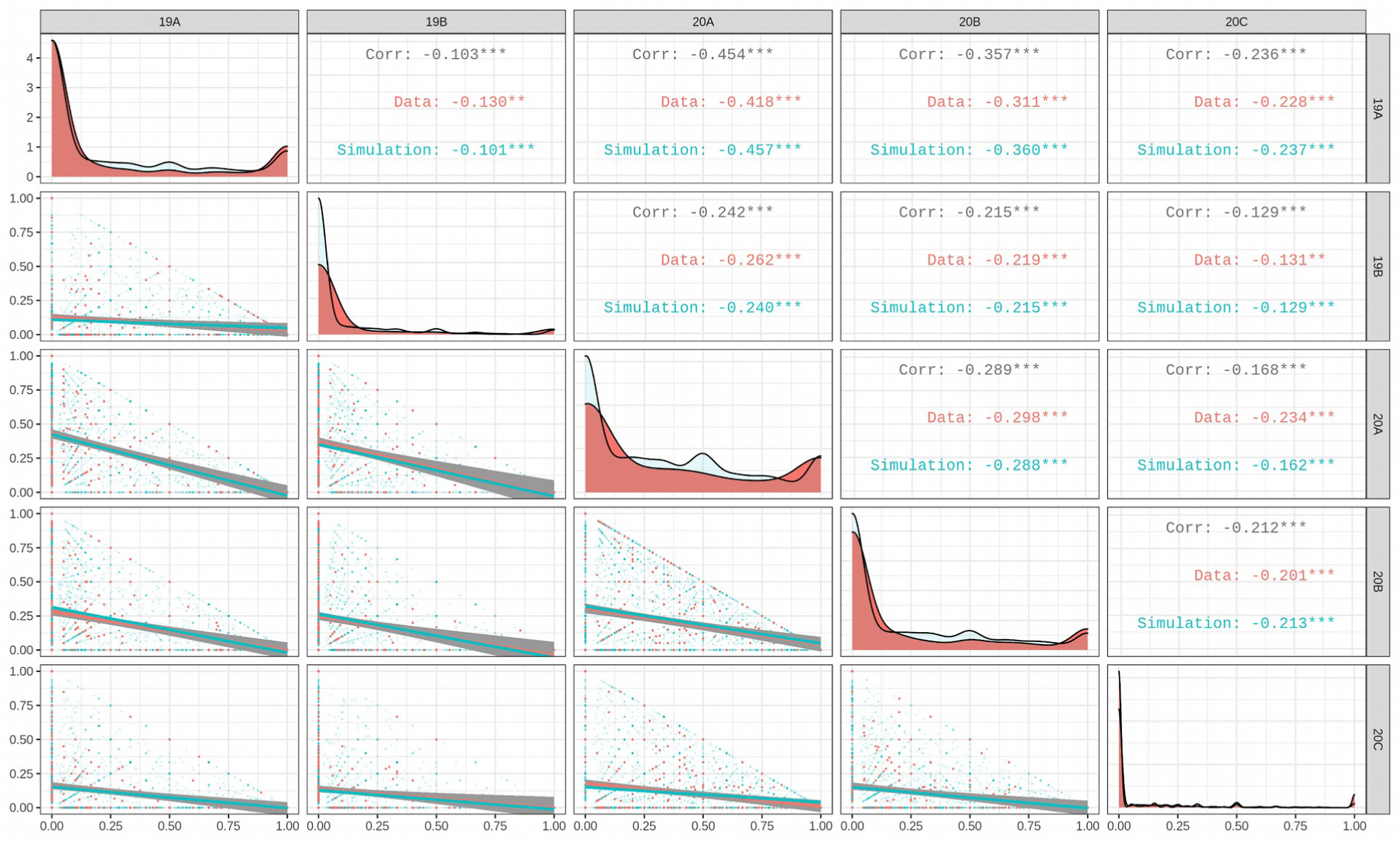
simulated (blue) against modeled data (red), showing the correlation, frequency and scatterplot of both plus the lines of the correlation estimation. As seen in the graph, the simulation is very similar to the data.

**Figure S2:**
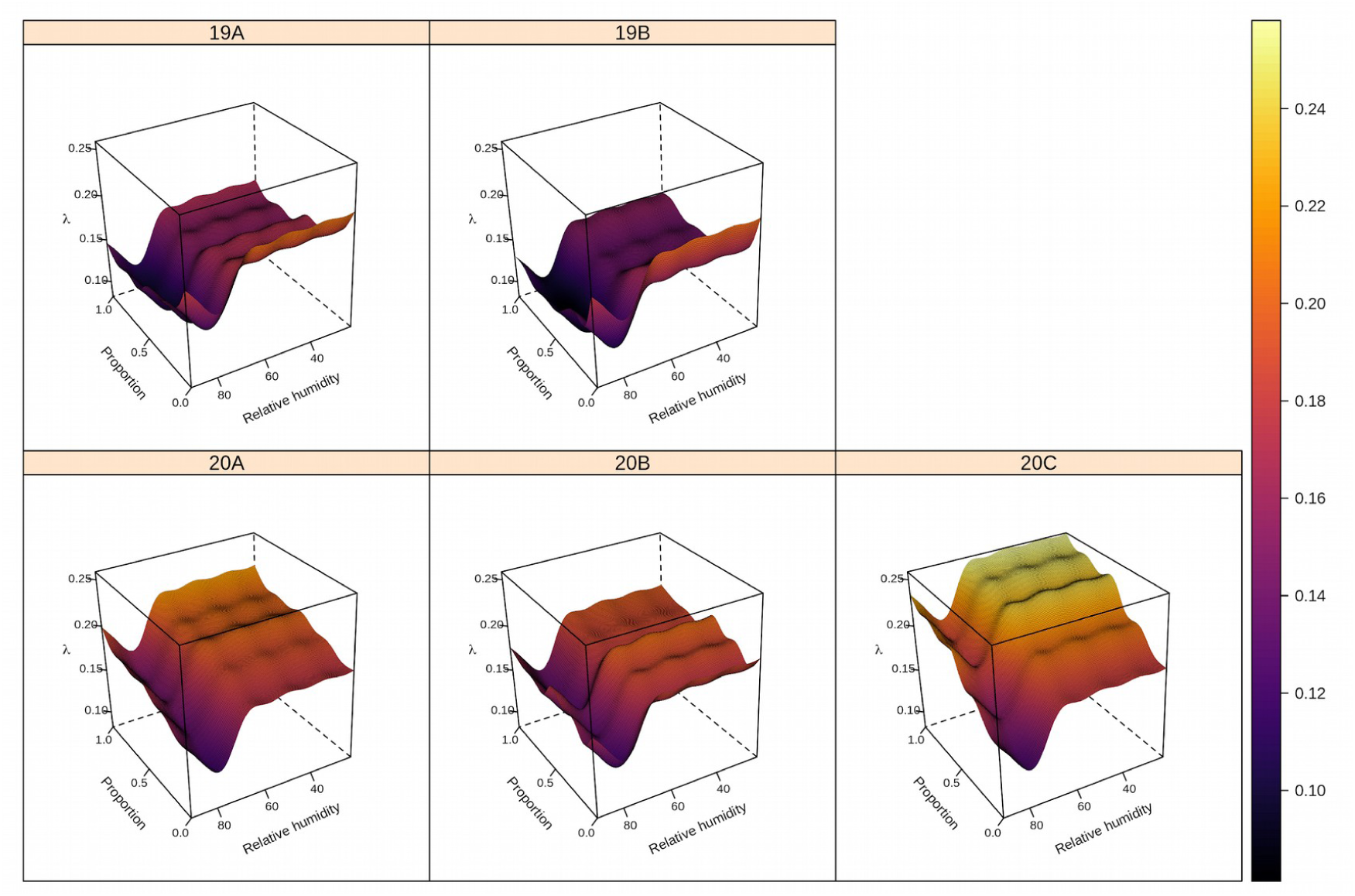
Predicted COVID-19 growth rate (color gradient from dark blue to yellow) based on the interaction between relative humidity and the proportion of clade 20C. Higher proportions of clade 20C and lower relative humidity are associated with higher values of COVID-19 growth rate. The clumps are a byproduct of GAM-smoothing across the discrete nature of classifications by nodes in Boosted Regression Trees.

**Figure S3.**
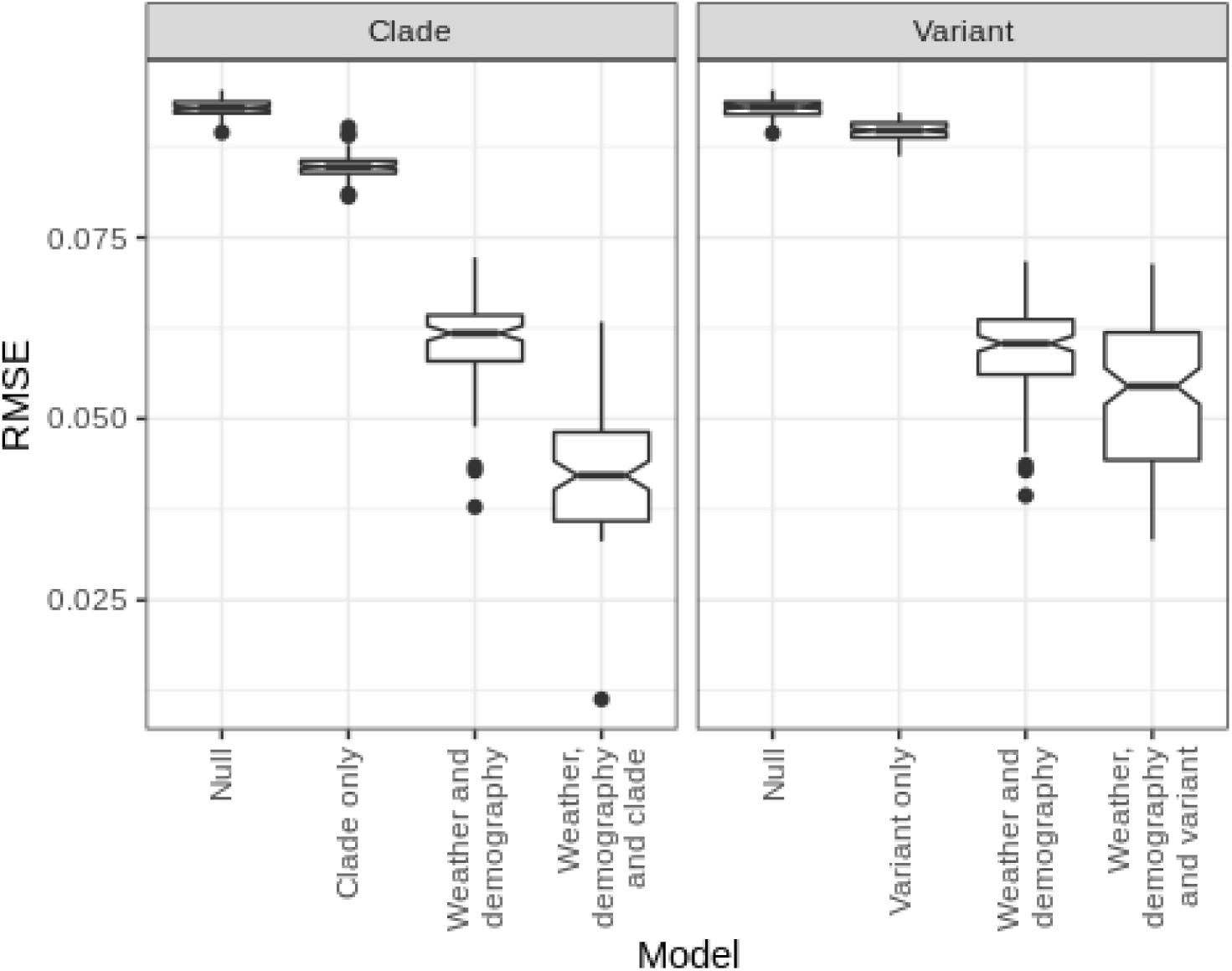
The root mean square error (RMSE) for all training sets, where lower RMSE indicates better performing models. Differences between null models and weather and demography models between the protein and mutation analyses arise because of different datasets for each of them. The best model among the training sets is the weather, demography and protein model.

**Figure S4.**
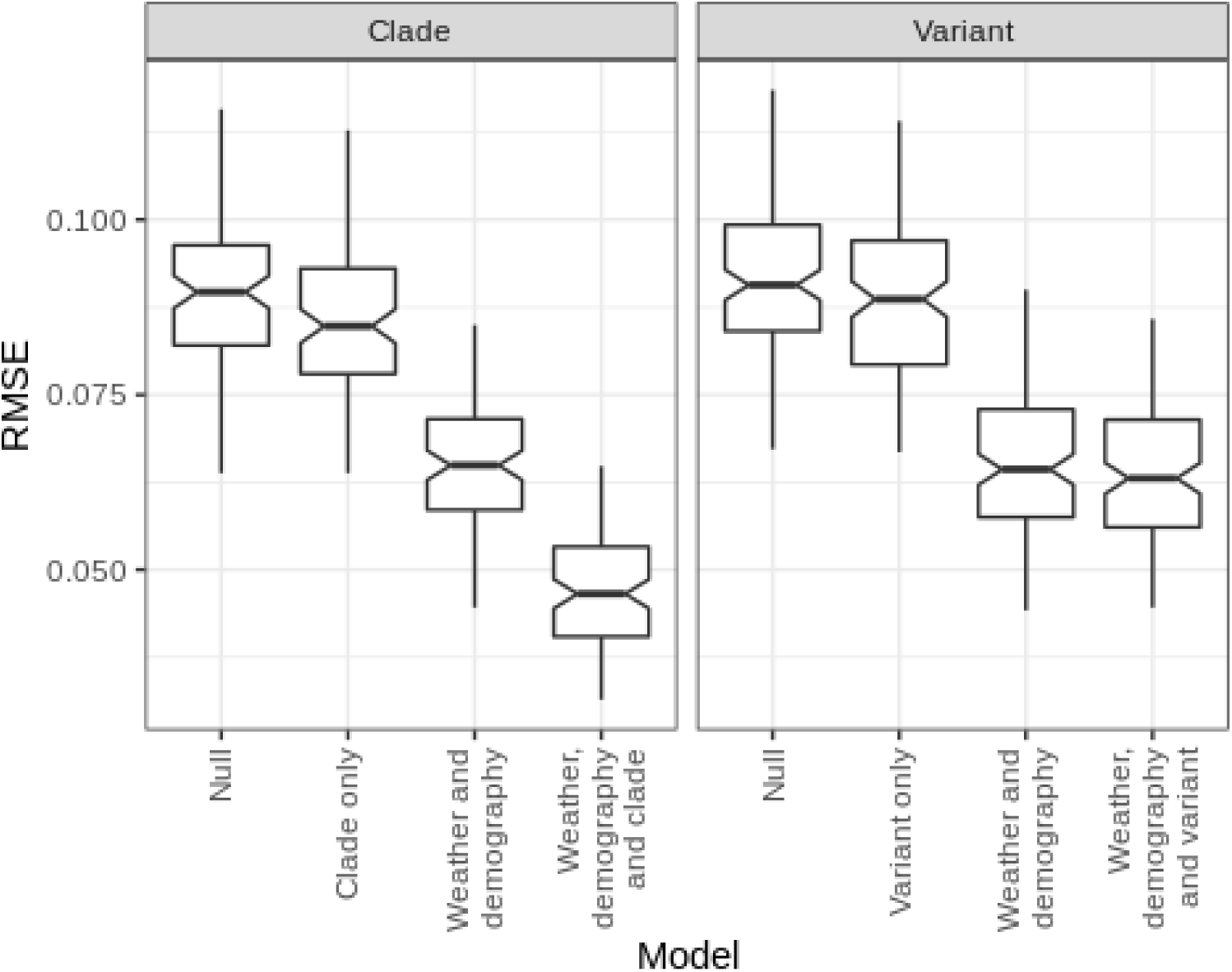
The RMSE for all testing sets. The null and weather and demography models differ because they are applied to different datasets consistent with either the mutation or protein data.The best model within the testing sets is the weather, demography and clade model.

**Figure S5.**
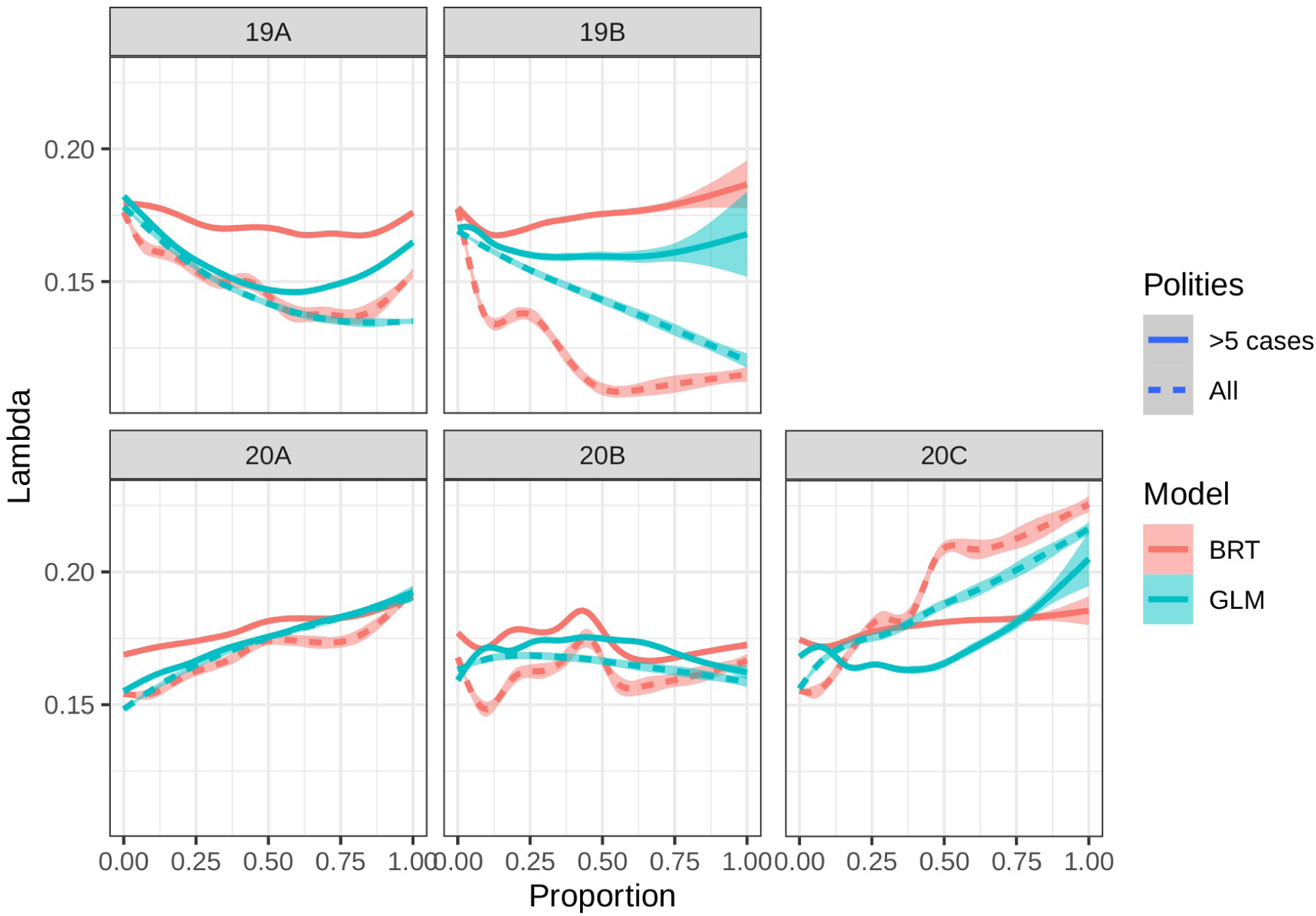
Sensitivity analysis to check if using only polities with five or more cases changes the results. This restriction reduced the number of polities from 319 to 248, losing nearly a fourth of the data. Despite the reductions in samples, the responses remained qualitatively similar, except that Clade 19 changes from a negative to a flat relationship. Both clades 20A and 20C maintain their positive relationship between prevalence and COVID-19 growth rate.

**Figure S6.**
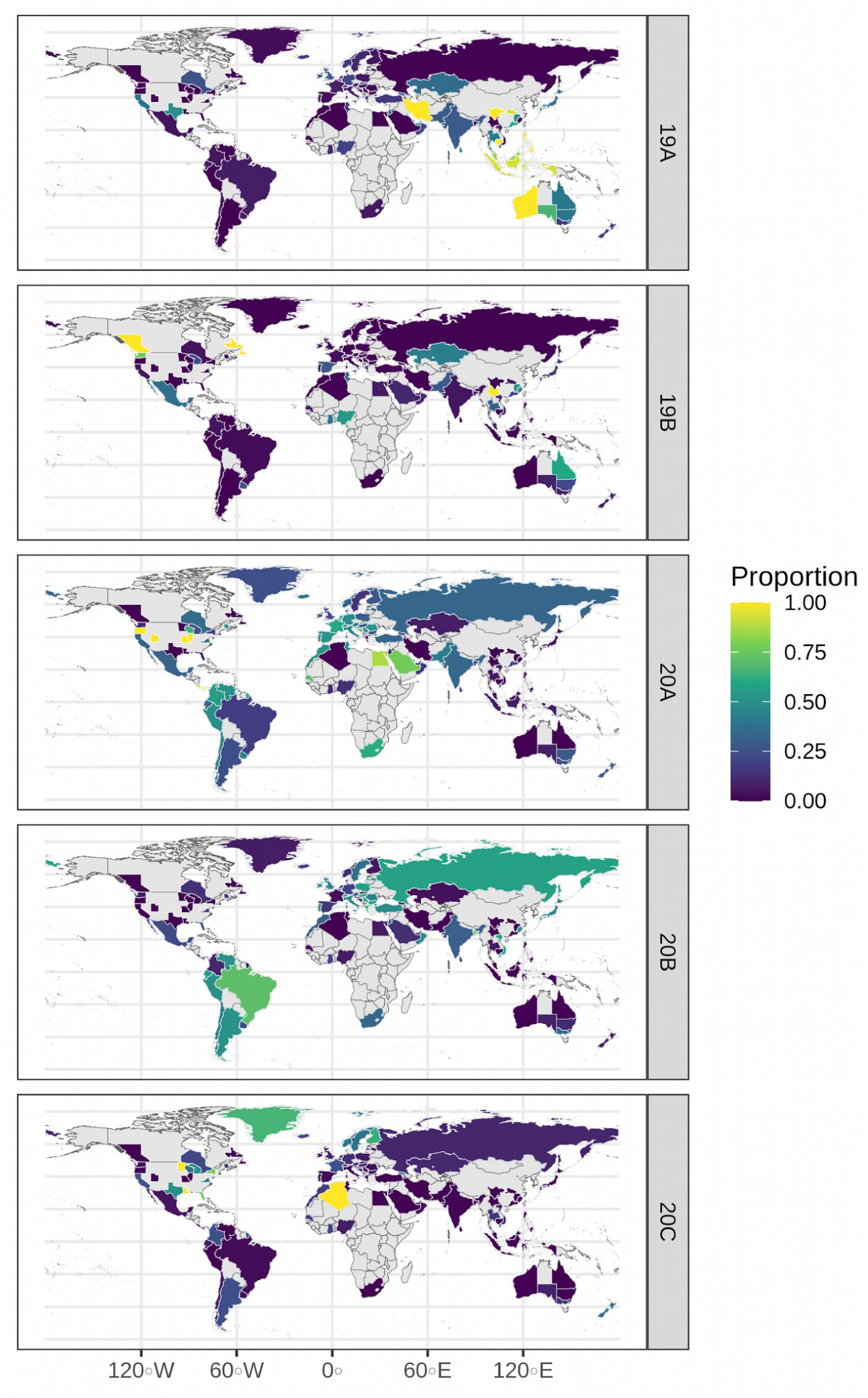
Proportions of each clade for every measured polity, as shown in the map clades 20B and 20A are more common in the americas, and europe, while 19A is most common.

**Figure S7.**
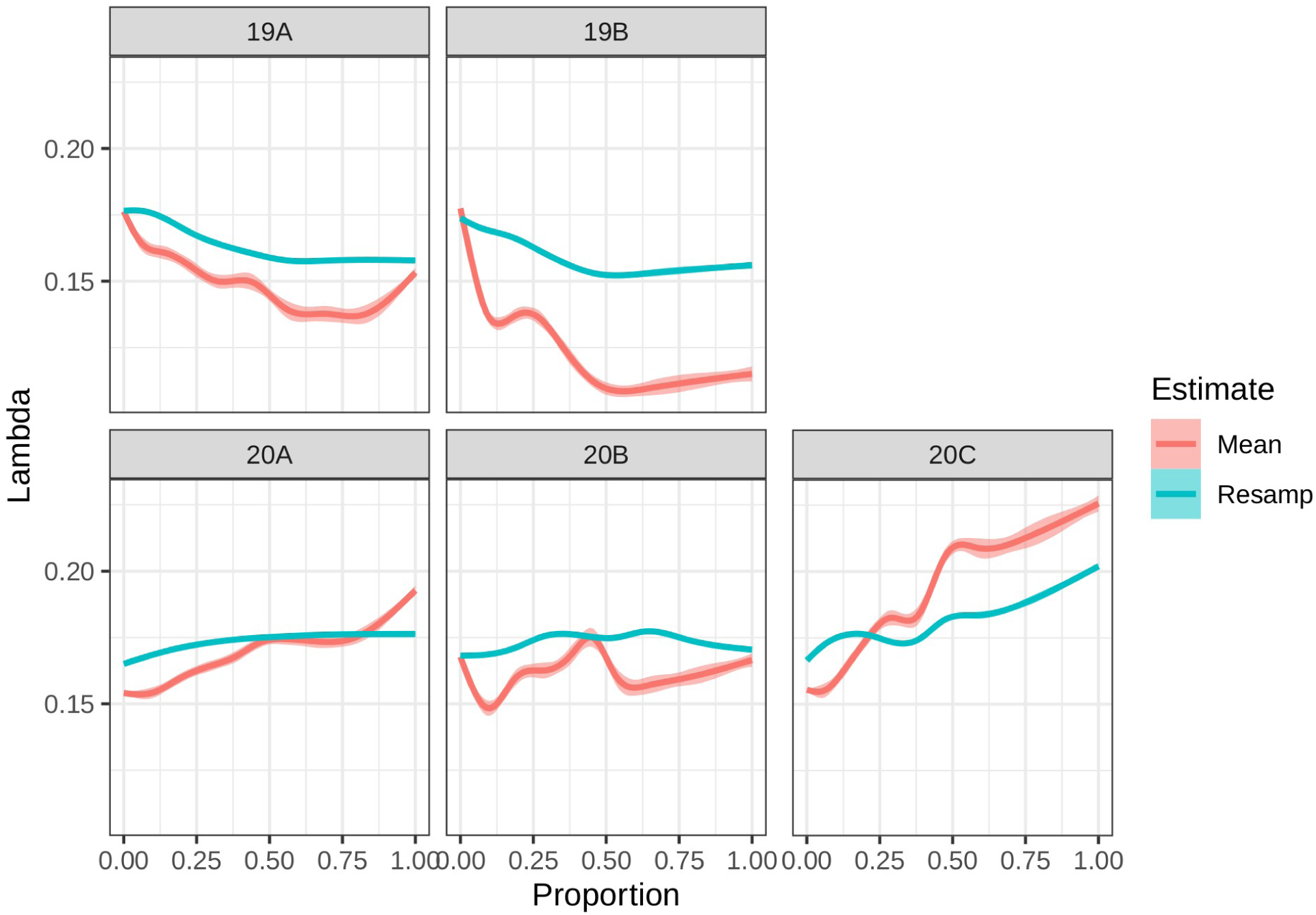
Sesnitivity analysis showing the response of remodeling with each polity resampled 100 tiemes choosing randomly between the Estimate, the upper and lower bound of a 95% multinomial confidence interval, the mean (red line), corresponds to the estimation using our models, while the resampled results are in blue. The only clade that seems to change is 19B which tends to have a flat response instead of having a negative relationship with Lambda.

**Table S1.**
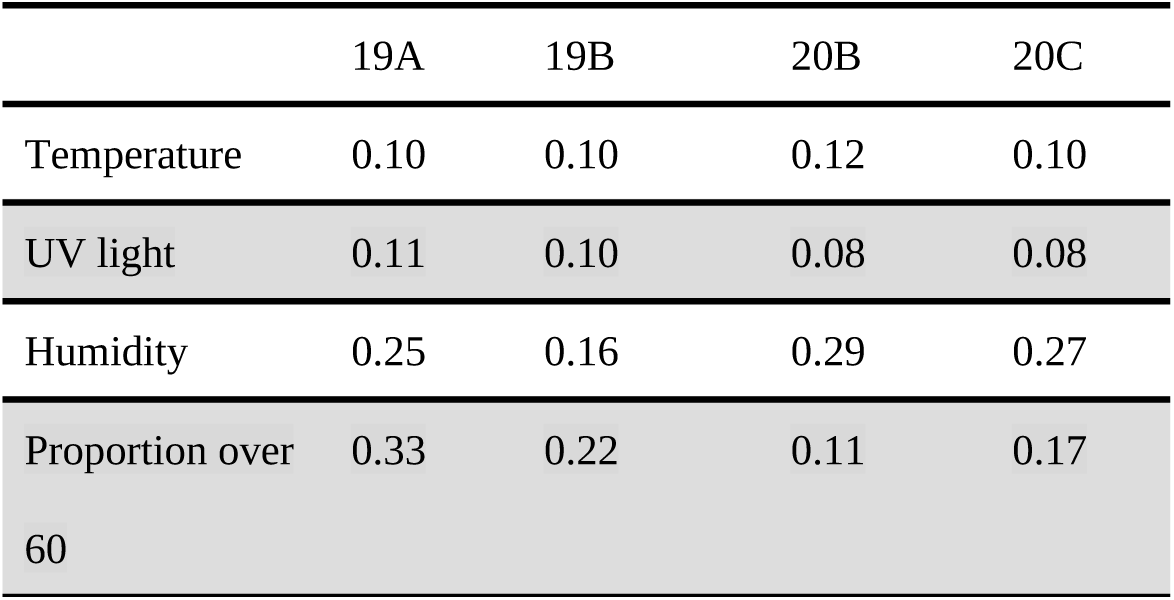
Friedmans H-statistic between clades and weather variables. The relative humidity is almost twice as important as any other variable in terms of its interaction with clade 20C

**Table S2:** Mutation D614G, the mutation which increased during the pandemic and has been suggested to be favored, is mostly found in type 2 clades, and very rarely in protein 19B. Most of the cases of mutation 614D correspond to clades of type 1, with only 5 cases corresponding to clade 20A.

| Variant\Clade | 19A | 19B | 20A | 20B | 20C | Sum |
| --- | --- | --- | --- | --- | --- | --- |
| **614D** | 611 | 221 | 5 | 0 | 0 | 837 |
| **614G** | 7 | 0 | 885 | 716 | 340 | 1948 |
| **614X** | 0 | 0 | 4 | 2 | 2 | 8 |
| **Sum** | 618 | 221 | 894 | 718 | 342 | 2793 |

## Data Availability

To be added to a github repository once the paper is accepted

